# Application accuracy of the sleep decision tree to standardized patient cases by physiotherapists: an observational study

**DOI:** 10.1101/2020.06.01.20119677

**Authors:** Catherine F. Siengsukon, Jianghua He, Kenneth Miller, Dianne Jewell

**Affiliations:** Department of Physical Therapy and Rehabilitation Science, University of Kansas Medical Center, Kansas City, KS; Department of Biostatistics & Data Science, University of Kansas Medical Center, Kansas City, KS; Department of Physical Therapy, University of North Texas Health Science Center, Fort Worth, TX; The Rehab Intelligence Network, Richmond, VA

**Keywords:** sleep decision tree, physiotherapy, patient cases

## Abstract

**Background:** Physiotherapists assess lifestyle factors, including sleep health, that contribute to poor health outcomes. Recommendations of sleep screening assessments have been provided; however, physical therapists’ ability to successfully identify which patients would benefit from additional consultation has not been established.

**Objective:** To determine if physiotherapists can accurately apply an evidence-based sleep decision tree to four hypothetical standardized patient cases.

**Methods:** Participants applied the sleep decision tree to the four standardized cases via an online platform. Likert scales were used to assess perception of ease of use, likelihood of use, and how helpful they thought the sleep decision tree would be. Descriptive analyses and multiple linear regression models were conducted.

**Results:** Eighty-eight individuals participated in the study. Eighty-one respondents (92%) correctly answered the least complex case while 12 (14%) correctly answered the most complex case. Seventy-four (84%) respondents indicated the sleep decision tree was easy to use, 57 (65%) answered they were likely to use the sleep decision tree in clinical practice, and 66 (75%) said the sleep decision tree would be helpful to their clients.

**Conclusions:** Physiotherapists were able to accurately apply a sleep decision tree to simpler patient cases but were frequently unable to apply it to more complex patient cases. This may be due to lack of education, perceived ease of using, and relevance of the sleep decision tree to their clinical practice. The sleep decision tree may aid physiotherapists in assessing sleep health, screening for sleep disturbances, and referring for further assessment.

## Introduction

Consistent adequate sleep quality is necessary for optimal function of the human body. Sleep disruption is associated with impaired pain modulation, tissue repair, immune function, physical and cognitive function, and endocrine function as well as an increased risk of cardiovascular disease, obesity, diabetes, depression, anxiety, mental health disorders, cancer, and Alzheimer’s disease.^1–10^ Sleep disturbances have also been associated with reduced physical activity and impaired physical function,^11,12^ which has direct implications for physiotherapists. An increased focus has been placed on identifying individuals who have a sleep disturbance or disorder and appropriately treating the sleep issue to enhance health and wellbeing and potentially delay or prevent the contribution of sleep disturbance to the development of chronic conditions.

Physiotherapists have been recognized as instrumental members of the health care team to assess lifestyle factors, including sleep health, that may contribute to poor health outcomes and could potentially be modified to enhance health outcomes.^13,14^ Furthermore, as physiotherapist become more integrated into primary care models^15,16^ and direct access continues to expand as it is found to be feasible, safe, improves patient outcomes and is cost-effective,^17,18^ physiotherapists have added responsibility to screen for a broader range of health conditions. Assessing sleep health and screening for sleep disturbances has been recommended as minimal health competencies^19^ and key skills for physiotherapists.^20^ Included in these recommendations is the referral of the individual to other health care professionals based on the results of these assessments.

A survey of physiotherapists’ perception and attitude about sleep showed that 82% of respondents agreed that physiotherapists should assess patients’ sleep quality and habits, and 95% agreed that addressing sleep issues may impact outcomes of physiotherapist services.^21^ However, a large majority of respondents (75%) reported they did not receive education about sleep during their entry-level education or since graduation (86%). The two most common reasons given for not routinely assessing their patients’ sleep quality or habits was “I do not know how to assess sleep habits or sleep quality” (25 of 48 responses) and “I do not have time” (7 of 48 responses). These results could indicate that physiotherapists need further education and skill development to efficiently and effectively assess their patients’ sleep health and screen for sleep disturbances.

Recent publications have been targeted to physiotherapists with recommendations for appropriate sleep screening methods and sleep health promotion techniques.^22,23^ However, guidance on when to refer the patient to another health care professional based on the results of the screening was not clearly delineated. A decision tree was developed as part of a sleep management tool kit for the American Physical Therapy Association’s Home Health Section with input from physiotherapists and sleep medicine experts.^24^ This evidence-based algorithm guides the physiotherapist to ask patients questions regarding their sleep health and sleep disturbances, to consider the clients’ current health condition that may be impacting their sleep health, to refer the client to a physician based on reliable and validated screening assessments, and to provide recommendations for sleep health promotion.

Appropriate use of this algorithm in clinical practice is untested. Therefore, the purpose of this study was to determine if physiotherapists were able to accurately apply the sleep decision tree to hypothetical standardized patient cases of varying complexity. A secondary purpose was to assess ease of use of sleep decision tree, likely of use in clinical practice the sleep decision tree, and perception on how helpful the sleep decision tree would be to clients in clinical practice.

## Materials and Methods

Four hypothetical patient cases were developed by the lead author. Each case described a scenario containing information about the patient’s presenting symptoms or diagnosis, past medical history, sleep duration and/or quality, as well as extraneous information that was not necessary for navigating the sleep decision tree but may be gathered as part of a typical physiotherapy examination. Each case had a distinct level of complexity that resulted in different decision and endpoints on the algorithm. The case with the least complexity required one decision within the algorithm. Another case required a decision at three points in the algorithm, and another required a decision at four points in the algorithm. The most complex case required a decision at five points in the algorithm.

The cases then underwent a two-stage process of revision to establish face validity of the cases. In the first stage, three experienced practicing physiotherapists reviewed the cases for clinical relatedness, similarity in difficulty, and adequate clinically reasonable distractors to ensure the correct answer was not obvious. Feedback was obtained through email or in-person or phone meeting, depending on the physiotherapist’s preference. The cases were then revised based on their feedback. In the next stage, the cases were reviewed by a psychologist who is certified in behavioral sleep medicine, a psychologist fellow, and a neurologist board certified in sleep medicine to ensure a reasonable level of difficulty considering the respondents would likely have a limited knowledge in sleep disturbances or screening for sleep disturbances and adequate distractors to ensure the correct answer was not obvious. The cases were again revised based on their feedback.

The finalized cases were loaded into REDCap (Research Electronic Data Capture), a secure, web-based data capture tool^25^ hosted at the (removed for masking). The cases were presented in a random order to avoid presentation in order of complexity. Branching logic was used to allow decisions to be made at various points within the algorithm as needed. Participants were advanced to the next screen and were not allowed to go back and change their answer once they submitted it. This study was conducted in accordance with the (removed for masking) Institutional Review Board.

The minimum sample size required was determined based on the power of the study in detecting that the rate of correct answer achieved using the decision-tree for a case is over 80%. Assuming the rate of correct answer achieved for a case is 90%, this study has 80% power of detecting the difference using a two-sided one sample z-test of a proportion against a known value (80%) with 83 participants.

Physiotherapists were invited to participate in this study through postings on the APTA’s Council on Prevention, Health Promotion, and Wellness in Physical Therapy community forum, the Academy of Prevention and Health Promotion Therapies community group, and email invitation to physiotherapists who are (removed for masking) alumni and clinical instructors. Individuals were asked to only participate if they were a physiotherapist.

Data were collected and managed using REDCap. Participants were asked to provide the following demographic information: sex, age, entry-level physiotherapist academic degree, how many years ago they graduated from entry-level physiotherapy program, current employment status as a physiotherapist providing direct patient care, the state(s) in which they are currently practicing, what age ranges(s) of patients they primary treat, and what type of setting(s) best describes their current primary work setting.

Participants were then asked to download the decision tree which was provided as a PDF. They were asked to “become familiar with the sleep decision tree” before starting to apply the algorithm to the four hypothetical patient cases. Participants also were asked how long they spent reading/reviewing the sleep decision tree before starting the cases. Participants then used a 10-point Likert scale to rate how easy or difficult it was to use the sleep decision tree with 1 being “very difficult” and 10 being “very easy”. They were asked on a 10-point Likert scale how likely they were to use the sleep decision tree in clinical practice with 1 being “not likely” to 10 being “very likely”. Lastly, they were asked on a 10-point Likert scale how helpful they thought the sleep decision tree would be to clients in their clinical practice with 1 being “not helpful” to 10 being “very helpful”.

Data were downloaded from REDCap and entered into IBM SPSS Statistics version 22. Means and standard deviations were calculated for continuous variables, and frequency distributions were calculated for categorical data. A two-sided one sample z-test of a proportion was conducted to determine if the proportion of respondents who answered each case correctly was statistically different than a known value (80%). Four multiple linear regression models were used to determine which factors explained the number of cases answered correctly, difficulty with using the sleep decision tree, likelihood that the respondent would use the sleep decision tree in their clinical practice, and how helpful the sleep decision tree would be to clients in their clinical practice. Five independent variables were chosen for each model as follows: sex, age, entry-level physiotherapy academic degree, number of years ago since graduation from entry-level physiotherapy education, and amount of time in minutes reading and/or reviewing the sleep decision tree before starting the cases. To reduce multicollinearity, age was removed from the models leaving four predictor variables in each model. Multicollinearity was assessed after age was removed and was below an acceptable level.

## Results

Of the 88 participants in this study, 65 (74%) were female, 60 (68%) received a doctorate for their entry-level physiotherapy academic degree, and 65 (74%) work full-time as a physiotherapy providing direct patient care (demographic details are found in Table 1). The average age of respondents was 40.8 (SD 10.9) years old. The largest number of respondents (n=23; 26%) graduated from entry-level physiotherapy education 1-5 years ago. The respondents reported treating patients from a variety of age ranges, with the largest number of respondents reporting treating primarily treat adults (>21 years old; n=122; 73%). Respondents work in a variety of setting with the largest number of respondents identifying their primary work setting as an outpatient clinic (n=40; 33%) followed by acute care/hospital (n=21; 15%). Respondents reported practicing in 22 different states, and the two states with the largest number of respondents currently practicing in Kansas (n=39; 41%) and Missouri (n=23; 24%). Respondents reported spending on average 4.7 minutes (SD 3.2; range 0-15 minutes) reading and/or reviewing the sleep decision tree before starting the cases.

**Table 1.** Demographics.

| Sex |  | Age<br>(years) | Which is your<br>entry-level<br>physiotherapy<br>academic<br>degree? |  | How many<br>years ago did<br>you graduate<br>from entry-level<br>physiotherapy<br>education? |  | What is your<br>current<br>employment<br>status as a<br>physiotherapy<br>providing<br>direct patient<br>care? |  | Which state(s) are<br>you currently<br>practicing? (more<br>than one response<br>permitted) |  | What age range(s) of<br>patients do you primarily<br>treat? (more than one<br>response permitted) |  | What type of setting best describes your<br>current primary work setting? (more than<br>one response permitted) |  |
| --- | --- | --- | --- | --- | --- | --- | --- | --- | --- | --- | --- | --- | --- | --- |
| N |  |  | N |  | N |  | N |  | N |  | N |  | N |  |
| 65 | F | 40.8 | 13 | Bachelor | 23 | 1-5 | 65 | Full time | 1 | Alaska | 8 | Birth to 3 yrs | 21 | Acute care/hospital |
| 23 | M | (10.9) | 15 | Master | 14 | 6-10 | 12 | Part time | 1 | Arkansas | 11 | 3yrs to school-age | 17 | Rehabilitation unit/facility |
|  |  |  | 60 | Doctorate | 11 | 11-15 | 9 | prn | 3 | California | 26 | School-age to 21 yrs | 3 | Sub-acute rehab |
|  |  |  |  |  | 12 | 16-20 | 2 | not | 2 | Colorado | 60 | 21-65 yrs | 40 | Outpatient clinic |
|  |  |  |  |  | 10 | 21-25 |  | providing | 1 | Florida | 62 | 65 yrs or older | 4 | School setting |
|  |  |  |  |  | 5 | 26-30 |  | direct | 1 | Georgia |  |  | 2 | Wellness/prevention/sports/fitness |
|  |  |  |  |  | 13 | >30 |  | patient care | 1 | Idaho |  |  | 6 | Extended care facility/nursing home/SNF |
|  |  |  |  |  |  |  |  |  | 4 | Illinois |  |  | 3 | Industrial/Workplace/Occupational<br>Environments |
|  |  |  |  |  |  |  |  |  | 39 | Kansas |  |  | 0 | Hospice |
|  |  |  |  |  |  |  |  |  | 1 | Maine |  |  | 3 | Local, State, and Federal Government |
|  |  |  |  |  |  |  |  |  | 1 | Mississippi |  |  | 1 | Research Center |
|  |  |  |  |  |  |  |  |  | 23 | Missouri |  |  | 10 | Faculty in physiotherapist or<br>physiotherapist assistant program |
|  |  |  |  |  |  |  |  |  | 1 | Montana |  |  | 0 | Not currently practicing as a clinician |
|  |  |  |  |  |  |  |  |  | 1 | Nebraska |  |  | 5 | Supervisor or administrator |
|  |  |  |  |  |  |  |  |  | 3 | New Jersey |  |  | 6 | Other |
|  |  |  |  |  |  |  |  |  | 1 | New Mexico |  |  |  |  |
|  |  |  |  |  |  |  |  |  | 1 | North Carolina |  |  |  |  |
|  |  |  |  |  |  |  |  |  | 1 | Tennessee |  |  |  |  |
|  |  |  |  |  |  |  |  |  | 3 | Texas |  |  |  |  |
|  |  |  |  |  |  |  |  |  | 2 | Utah |  |  |  |  |
|  |  |  |  |  |  |  |  |  | 2 | Virginia |  |  |  |  |
|  |  |  |  |  |  |  |  |  | 1 | Washington |  |  |  |  |

Eighty-one respondents (92%; p=0.002) correctly answered the least complex case with one decision point, and 74 (84%) correctly answered the case with three decision points in the algorithm (Table 2). For the more complex cases, 6 (7%) correctly answered the case with four decision points, and 12 (14%) correctly answered the case with five decision points in the algorithm (Table 3). One (1%) respondent answered none of the cases correctly, 17 (19%) answered one of the cases correctly, 55 (63%) answered two of the cases correctly, 14 (16%) answered three of the cases correctly, and one (1%) respondent answered all four cases correctly (Table 3).

**Table 2.** Number of participants who correctly answered the case by case

| **Level of Complexity (number of decisions required to answer case correctly)** | **Number of participants who correctly answered the case (n; %)** | **Z-test p-value** |
| --- | --- | --- |
| 1 | 81 (92) | 0.002 |
| 3 | 74 (84) | 0.169 |
| 4 | 6 (7) | 1 |
| 5 | 12 (14) | 1 |

**Table 3.** Number of cases participants answered correctly.

| **Number of Cases Answered**<br>**Correctly** | **Number of participants**<br>**(n; %)** |
| --- | --- |
| 0 | 1 (1) |
| 1 | 17 (19) |
| 2 | 55 (63) |
| 3 | 14 (16) |
| 4 | 1 (1) |

There were a range of responses given on the Likert-scales assessing difficulty/ease with using the sleep decision tree, likelihood that the respondent would use the sleep decision tree in their clinical practice, and how helpful the sleep decision tree would be to clients in their clinical practice (Table 4). The highest number of respondents (n=21; 24%) answered that the sleep decision tree was “very easy” to use. If we include all respondents who answered ≥6 on the Likert-scale, 74 (84%) respondents indicated the sleep decision tree was easy to use. Ten (11%) respondents answered that they were “not likely” to use the sleep decision tree in their clinical practice, and 10 (11%) answered that they were “very likely” to use the sleep decision tree in their clinical practice. If we include all respondents who answered ≥6 on the Likert-scale, 57 (65%) respondents indicated they were likely to use the sleep decision tree in clinical practice. Six (7%) respondents answered that the sleep decision tree would not be helpful to their clients, and seventeen (19%) answered that it would be very helpful to their clients. If we include all respondents who answered ≥6 on the Likert-scale, 66 (75%) respondents indicated the sleep decision tree would be helpful to their clients.

**Table 4.** Likert-scales assessing level of difficulty in using the sleep decision tree, likelihood respondent would use the sleep decision tree in their clinical practice, and how helpful the sleep decision tree will be to clients in their clinical practice

| On a scale from 1 to 10 with 1 being very difficult to use to 10 being very easy to use, how easy or difficult was the sleep decision tree to use? |  |  |  |  |  |  |  |  |  |  |
| --- | --- | --- | --- | --- | --- | --- | --- | --- | --- | --- |
|  | Very Difficult<br>1 |  |  |  |  |  |  |  |  | Very Easy<br>10 |
| n (%) | 0 | 1 (1) | 3 (3) | 5 (6) | 5 (6) | 6 (7) | 17 (19) | 16 (18) | 14 (16) | 21 (24) |
| On a scale from 1 to 10 with 1 being not likely to use at all to 10 being very likely to use, how likely are you to use the sleep decision tree in clinical practice? |  |  |  |  |  |  |  |  |  |  |
|  | Not Likely<br>1 |  |  |  |  |  |  |  |  | Very Likely<br>10 |
| n (%) | 10 (11) | 0 (0) | 4 (5) | 6 (7) | 11 (13) | 10 (11) | 16 (18) | 15 (17) | 6 (7) | 10 (11) |
| On a scale from 1 to 10 with 1 being not helpful at all to 10 being very helpful, helpful do you think using the sleep decision tree will be to clients in your clinical practice? |  |  |  |  |  |  |  |  |  |  |
|  | Not Helpful<br>1 |  |  |  |  |  |  |  |  | Very Helpful<br>10 |
| n (%) | 6 (7) | 2 (2) | 3 (3) | 3 (3) | 8 (9) | 6 (7) | 20 (23) | 15 (17) | 8 (9) | 17 (19) |

The multiple linear regression models to determine which factors explained the number of cases answered correctly, difficulty with using the sleep decision tree, likelihood that the respondent would use the sleep decision tree in their clinical practice, and how helpful the sleep decision tree would be to clients in their clinical practice were not statistically significant (Table 5). In the first model of predicting the number of cases answered correctly, none of the variables had a significant effect on the model, although 6-10 years of practice was at p=0.055, that individuals practicing 6-10 years answered on average .459 cases more incorrectly compared to the group practicing 1-5 years. In the second model predicting the difficulty/ease with using the sleep decision tree, having an entry-level master’s degree was a significant predictor (p=0.033) indicating those with a master’s degree viewed the sleep decision tree on average 1.541 points more easy to use than those with an entry-level doctorate degree. Although not statically significant (p=0.089), individuals practicing for 30+ years reported on average 2.017 higher degree of ease of use than those practicing 1-5 years. In the third model predicting how likely participants were to use the sleep decision tree, men were 1.350 points less likely to use the sleep decision tree than women respondents (p=0.048). Although not statistically significant (p=0.098), those with a master’s degree were 1.563 points less likely to use the decision tree than those with a doctorate degree. In the third model predicting response to the question of how helpful the sleep decision tree would be to clients in their clinical practice, none of the variables had a significant effect on the model, although 6-10 years of practice was approaching statistical significance at p=0.055, that individuals practicing 6-10 years answered on average 1.721 points lower that the sleep decision tree would be helpful compared to the group practicing 1-5 years.

**Table 5.** Regression models.

| | B | SE | $\beta$ | t | Sig. (p) | R<sup>2</sup> |
| --- | --- | --- | --- | --- | --- | --- |
| **Model 1: Number of Correct Cases** |  |  |  |  | .592 | .098 |
| Sex (female reference) | .158 | .176 | .104 | .900 | .371 |  |
| Masters (Doctorate reference) | .329 | .243 | .186 | 1.351 | .181 |  |
| Bachelors (Doctorate reference) | -.032 | .366 | -.017 | -.086 | .931 |  |
| 6-10 yrs practice (1-5 yrs practice reference) | -.459 | .235 | -.252 | -1.949 | .055 |  |
| 11-15 yrs practice (1-5 yrs practice reference) | -.164 | .253 | -.081 | -.648 | .519 |  |
| 16-20 yrs practice (1-5 yrs practice reference) | -.376 | .271 | -.194 | -1.386 | .170 |  |
| 21-25 yrs practice (1-5 yrs practice reference) | -.385 | .296 | -.184 | -1.300 | .197 |  |
| 26-30 yrs practice (1-5 yrs practice reference) | -.255 | .346 | -.089 | -.737 | .463 |  |
| 30+ yrs practice (1-5 yrs practice reference) | -.415 | .389 | -.222 | -1.066 | .290 |  |
| Length of Time Reviewed Decision Tree | -.014 | .023 | -.066 | -.591 | .556 |  |
| **Model 2: Difficulty/Ease with Using the Sleep Decision Tree** |  |  |  |  | .342 | .129 |
| Sex (female reference) | -.194 | .528 | -.042 | -.367 | .715 |  |
| Masters (Doctorate reference) | 1.541 | .732 | .284 | 2.104 | .039 |  |
| Bachelors (Doctorate reference) | -1.639 | 1.101 | -.286 | -1.489 | .140 |  |
| 6-10 yrs practice (1-5 yrs practice reference) | -.177 | .709 | -.032 | -.250 | .803 |  |
| 11-15 yrs practice (1-5 yrs practice reference) | -1.042 | .761 | -.169 | -1.370 | .175 |  |
| 16-20 yrs practice (1-5 yrs practice reference) | -1.188 | .817 | -.200 | -1.453 | .150 |  |
| 21-25 yrs practice (1-5 yrs practice reference) | -1.355 | .891 | -.211 | -1.521 | .132 |  |
| 26-30 yrs practice (1-5 yrs practice reference) | -.253 | 1.041 | -.029 | -.243 | .809 |  |
| 30+ yrs practice (1-5 yrs practice reference) | 2.017 | 1.172 | .351 | 1.720 | .089 |  |
| Length of Time Reviewed Decision Tree | -.064 | .069 | -.103 | -.935 | .353 |  |
| **Model 3: How Likely Use the Sleep Decision Tree** |  |  |  |  | .212 | .151 |
| Sex (female reference) | -1.350 | .673 | -.226 | -2.007 | .048 |  |
| Masters (Doctorate reference) | -1.563 | .933 | -.224 | -1.675 | .098 |  |
| Bachelors (Doctorate reference) | 1.347 | 1.402 | .182 | .961 | .340 |  |
| 6-10 yrs practice (1-5 yrs practice reference) | -.990 | .902 | -.138 | -1.097 | .276 |  |
| 11-15 yrs practice (1-5 yrs practice reference) | -.918 | .969 | -.116 | -.947 | .346 |  |
| 16-20 yrs practice (1-5 yrs practice reference) | .408 | 1.041 | .053 | .392 | .696 |  |
| 21-25 yrs practice (1-5 yrs practice reference) | .519 | 1.135 | .063 | .457 | .649 |  |
| 26-30 yrs practice (1-5 yrs practice reference) | .872 | 1.325 | .077 | .658 | .513 |  |
| 30+ yrs practice (1-5 yrs practice reference) | -.921 | 1.493 | -.124 | -.617 | .539 |  |
| Length of Time Reviewed Decision Tree | .146 | .088 | .181 | 1.663 | .100 |  |
| **Model 4: How Helpful the Sleep Decision Tree Would be to Clients in Their Clinical Practice** |  |  |  |  | .210 | .151 |
| Sex (female reference) | -1.052 | .654 | -.181 | -1.609 | .112 |  |
| Masters (Doctorate reference) | -1.200 | .906 | -.177 | -1.324 | .189 |  |
| Bachelors (Doctorate reference) | 1.956 | 1.362 | .272 | 1.436 | .155 |  |
| 6-10 yrs practice (1-5 yrs practice reference) | -1.721 | .877 | -.247 | -1.963 | .053 |  |
| 11-15 yrs practice (1-5 yrs practice reference) | -1.336 | .941 | -.173 | -1.420 | .160 |  |
| 16-20 yrs practice (1-5 yrs practice reference) | .016 | 1.011 | .002 | .016 | .987 |  |
| 21-25 yrs practice (1-5 yrs practice reference) | -.364 | 1.103 | -.045 | -.330 | .742 |  |
| 26-30 yrs practice (1-5 yrs practice reference) | -.297 | 1.288 | -.027 | -.230 | .818 |  |
| 30+ yrs practice (1-5 yrs practice reference) | -2.331 | 1.451 | -.324 | -1.607 | .112 |  |
| Length of Time Reviewed Decision Tree | .111 | .085 | .141 | 1.302 | .197 |  |

## Discussion

This is the first study to determine if physiotherapists were able to accurately apply a sleep decision tree to patient cases. The results of this study demonstrate that physiotherapists were able to accurately apply a sleep decision tree to simpler patient cases but were frequently unable to apply the sleep decision tree to more complex patient cases. This may be due to lack of education on use of the sleep decision tree, perceived ease or difficulty with using the sleep decision tree, perceived relevance of the sleep decision tree to their clinical practice, or the cases were too vague.

While the majority of respondents in this study were able to accurately apply the sleep decision tree to less complex patient scenarios, few were able to accurately apply the sleep decision tree to more complex patient cases. It appears that number of years of practice may influence the number of cases answered correctly, with participants practicing for ≥6 years answering less cases correctly (not statistically significant) than participants practicing for 1-5 years. Perhaps this is because entry-level physiotherapy programs are integrating information about sleep and sleep health into their curriculum. Future studies are needed to determine if education of physiotherapists on use and application of the sleep decision tree would enhance ability to correctly apply the sleep decision tree to more complex cases. Study of the type, mode, and amount of education needed to optimize correct application of the sleep decision tree would be informative.

Surprisingly, more participants (n=12) answered the most complex case (5 decision points) correctly than the case with 4 decision points (n=6). In reviewing the cases, it is possible that it was not clear the patient in the case had a sleep issue prior to the reason he initiated physiotherapy services, which could have led the respondent to the incorrect response. Also, in the 4-decision and 5-decision point cases, respondents had to select two correct responses at one of the decision points. Even though the question stated to “select all that apply”, it appears that this was the decision point the majority of respondents answered incorrectly. It seems plausible that instructions prior to using the sleep decision tree would likely improve accuracy in applying the decision tree to cases. Also, applying the decision tree to actual patients rather than standardized cases might improve accuracy because clarifying questions could be asked.

Interestingly, participants with a master’s degree reported the sleep decision tree was easier to use than participants with a doctorate degree but were less likely (not statistically significant) to use the sleep decision tree in practice. Perhaps this is because individuals with a master’s degree have more exposure to decision trees in general but rely on them less due to practice expertise. It may also be due to lower sample size of participants with a master’s or bachelor’s degree. Also interesting, was that men reported they were less likely to use the sleep decision tree than females. This may also be due to the skewed distribution of male vs female participants. We did not include practice setting in the model because participants were able to select more than one response, but the sleep decision tree was developed based on best practice and valid and reliable cut-offs for adults, so would presumably not be helpful to pediatric patients or likely to be used in pediatric practice.

One limitation of this study is a majority of respondents primary practice in midwestern states. This may have impacted the results with respondents having access to continuing education courses and perhaps more integration of sleep health education into entry-level programs in midwestern states. Another limitation is the study may have been underpowered due to overestimating the rate of correct answers. While we enrolled a sufficient number of participants to meet the predetermined sample size, the rate of correct answers for the simplest case (one decision point) is 92% while those for more complicated cases were 84% (three decision points), 7% (four decision points), and 14% (five decision points), respectively. Therefore, this study may have adequate power for the simplest case but not for the more complicated cases.

With evidence demonstrating adequate quality sleep is critical for optimal health and to reduce the risk of mortality and emphasis of physiotherapists being part of health care team equipped to screen for sleep disturbances, this study was a necessary step in translating the recommendation for physiotherapists to screen for sleep disturbances into practice. Additional study is needed to understand how to optimize application of the sleep decision tree, and then steps are needed to determine how to translate the sleep decision tree into clinical practice.

**Figure 1.**
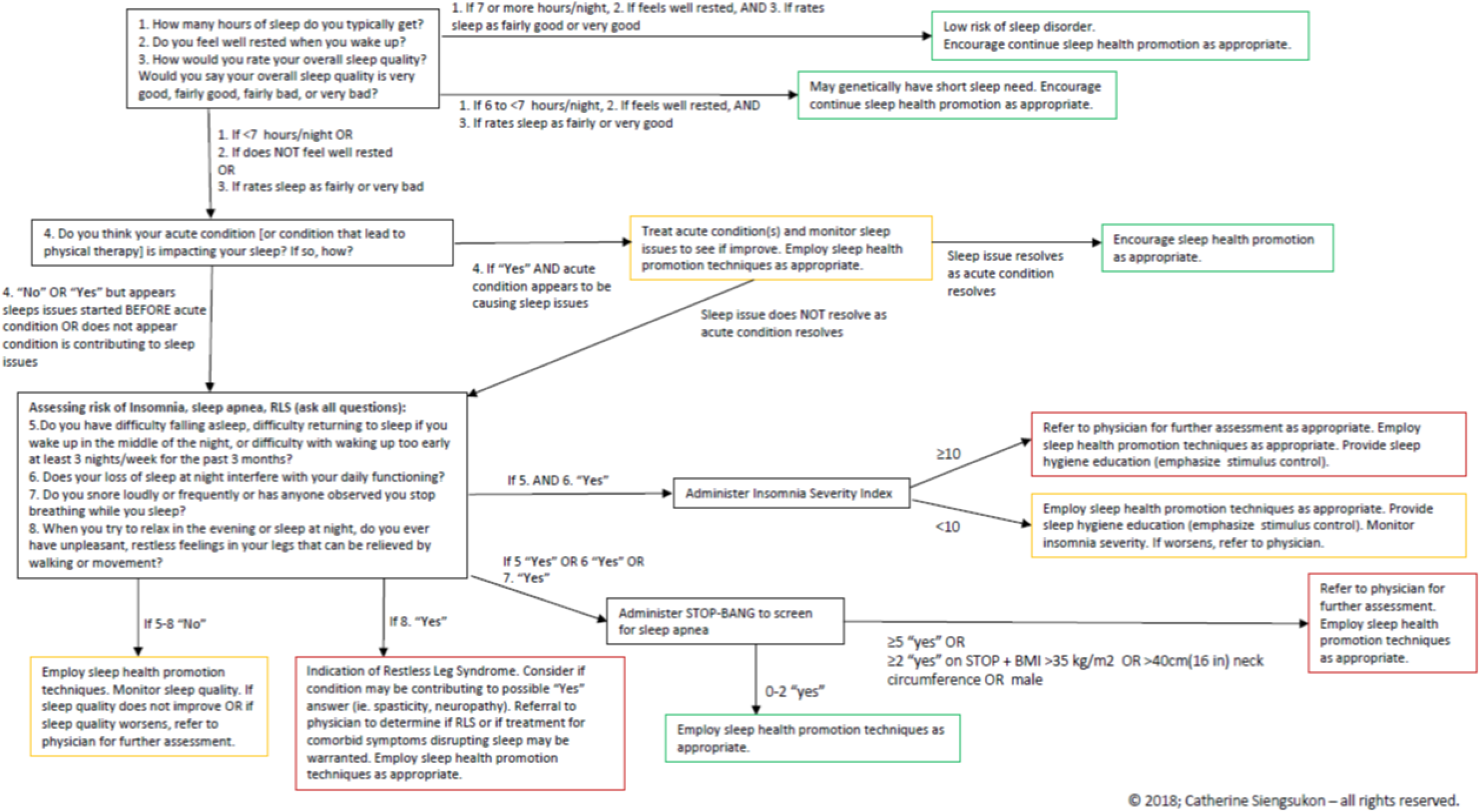
Sleep decision tree

## Data Availability

Data will be available upon request to corresponding author

## Acknowledgements

We would like to acknowledge Stacia Troshynski Brown, Alexandra Bea, Michelle Drerup, and Suzanne Stevens for their contribution to reviewing and providing feedback on the cases.

## References

1. Research CoSMa. Sleep Disorders and Sleep Deprivation: An Unmet Public Health Problem. Washington DC: National Academies Press; 2006.

2. Besedovsky L, Lange T, Born J. Sleep and immune function. Pflugers Arch. 2012;463(1):121–137.

3. Finan PH, Goodin BR, Smith MT. The association of sleep and pain: an update and a path forward. J Pain. 2013;14(12):1539–1552.

4. Aziz M, Ali SS, Das S, et al. Association of Subjective and Objective Sleep Duration as well as Sleep Quality with Non-Invasive Markers of Sub-Clinical Cardiovascular Disease (CVD): A Systematic Review. J Atheroscler Thromb. 2017;24(3):208–226.

5. St-Onge MP, Grandner MA, Brown D, et al. Sleep Duration and Quality: Impact on Lifestyle Behaviors and Cardiometabolic Health: A Scientific Statement From the American Heart Association. Circulation. 2016;134(18):e367-e386.

6. Cappuccio FP, D’Elia L, Strazzullo P, Miller MA. Sleep duration and all-cause mortality: a systematic review and meta-analysis of prospective studies. Sleep. 2010;33(5):585–592.

7. Taylor DJ, Lichstein KL, Durrence HH, Reidel BW, Bush AJ. Epidemiology of insomnia, depression, and anxiety. Sleep. 2005;28(11):1457–1464.

8. Taylor DJ, Mallory LJ, Lichstein KL, Durrence HH, Riedel BW, Bush AJ. Comorbidity of chronic insomnia with medical problems. Sleep. 2007;30(2):213–218.

9. Brown BM, Rainey-Smith SR, Villemagne VL, et al. The Relationship between Sleep Quality and Brain Amyloid Burden. Sleep. 2016;39(5):1063–1068.

10. Ju YE, Lucey BP, Holtzman DM. Sleep and Alzheimer disease pathology--a bidirectional relationship. Nat Rev Neurol. 2014;10(2):115–119.

11. Kyle SD, Morgan K, Espie CA. Insomnia and health-related quality of life. Sleep medicine reviews. 2010;14:69–82.

12. Kline CE. The bidirectional relationship between exercise and sleep: Implications for exercise adherence and sleep improvement. Am J Lifestyle Med. 2014;8(6):375–379.

13. Dean E. Physical therapy in the 21st century (Part I): toward practice informed by epidemiology and the crisis of lifestyle conditions. Physiother Theory Pract. 2009;25(5–6):330–353.

14. Dean E. Physical therapy in the 21st century (Part II): evidence-based practice within the context of evidence-informed practice. Physiother Theory Pract. 2009;25(5–6):354–368.

15. Murphy BP, Greathouse D, Matsui I. Primary care physical therapy practice models. J Orthop Sports Phys Ther. 2005;35(11):699–707.

16. Maharaj S, Chung C, Dhugge I, et al. Integrating Physiotherapists into Primary Health Care Organizations: The Physiotherapists’ Perspective. Physiother Can. 2018;70(2):188–195.

17. Ojha HA, Snyder RS, Davenport TE. Direct access compared with referred physical therapy episodes of care: a systematic review. Phys Ther. 2014;94(1):14–30.

18. Piscitelli D, Furmanek MP, Meroni R, De Caro W, Pellicciari L. Direct access in physical therapy: a systematic review. Clin Ter. 2018;169(5):e249-e260.

19. Dean E, Greig A, Murphy S, et al. Raising the Priority of Lifestyle-Related Noncommunicable Diseases in Physical Therapy Curricula. Phys Ther. 2016;96(7):940–948.

20. Bezner JR. Promoting Health and Wellness: Implications for Physical Therapist Practice. Phys Ther. 2015;95(10):1433–1444.

21. Siengsukon CF, Al-Dughmi M, Sharma NK. A survey of physical therapists’ perception and attitude about sleep. Journal of allied health. 2015;44(1):41–50.

22. Siengsukon CF, Al-Dughmi M, Stevens S. Sleep Health Promotion: Practical Information for Physical Therapists. Phys Ther. 2017;97(8):826–836.

23. Nijs J, Mairesse O, Neu D, et al. Sleep Disturbances in Chronic Pain: Neurobiology, Assessment, and Treatment in Physical Therapist Practice. Phys Ther. 2018;98(5):325–335.

24. Siengsukon C, Miller KL. Sleep Management in the Home. 2018; https://www.homehealthsection.org/assets/docs/Sleep_Kit_3-2018.pdf. Accessed 11/8/2019.

25. Harris PA, Taylor R, Thielke R, Payne J, Gonzalez N, Conde JG. Research electronic data capture (REDCap)--a metadata-driven methodology and workflow process for providing translational research informatics support. J Biomed Inform. 2009;42(2):377–381.

